# BNT162b2 XBB1.5-adapted Vaccine and COVID-19 Hospital Admissions and Ambulatory Visits in US Adults

**DOI:** 10.1101/2023.12.24.23300512

**Authors:** Sara Y. Tartof, Jeff M. Slezak, Timothy B. Frankland, Laura Puzniak, Vennis Hong, Bradley K. Ackerson, Julie A Stern, Sarah Simmons, Luis Jodar, John M. McLaughlin

**Affiliations:** Kaiser Permanente Southern California Department of Research & Evaluation, Pasadena, CA, USA; Department of Health Systems Science, Kaiser Permanente Bernard J. Tyson School of Medicine, Pasadena, CA, USA; Kaiser Permanente Hawaii Center for Integrated Health Care Research, Honolulu, HI; Pfizer Inc., New York, NY, USA; Southern California Permanente Medical Group, Harbor City, CA, USA

## Abstract

**Importance:** Data describing the early additional protection afforded by recently recommended XBB1.5- adapted COVID-19 vaccines are limited.

**Objective:** We estimated the association between receipt of BNT162b2 XBB1.5-adapted vaccine (Pfizer-BioNTech 2023–2024 formulation) and medically attended COVID-19 outcomes among adults ≥18 years of age.

**Design, Setting, and Participants:** We performed a test-negative case-control study to compare the odds of BNT162b2 XBB1.5- adapted vaccine receipt between COVID-19 cases and test-negative controls among adults in the Kaiser Permanente Southern California health system between October 11 and December 10, 2023. Adjusted odds ratios (OR) and 95% confidence intervals (CI) were estimated from multivariable logistic regression models that were adjusted for patient demographic and clinical characteristics.

**Exposure:** The primary exposure was receipt of BNT162b2 XBB1.5-adapted vaccine compared to not receiving an XBB1.5-adapted vaccine of any kind, regardless of prior COVID-19 vaccination or SARS-CoV-2 infection history. We also compared receipt of prior (non-XBB1.5-adapted) versions of COVID-19 vaccines to the unvaccinated to estimate remaining protection from older vaccines.

**Main Outcomes and Measures:** Cases were those with a positive SARS-CoV-2 polymerase chain reaction test, and controls tested negative. Analyses were done separately for COVID-19 hospital admissions, emergency department (ED) and urgent care (UC) encounters, and outpatient visits.

**Results:** Among 4232 cases and 19,775 controls with median age of 54 years, adjusted ORs for testing positive for SARS-CoV-2 among those who received BNT162b2 XBB1.5-adapted vaccine a median of 30 days ago (*vs* not having received an XBB1.5-adapted vaccine of any kind) were 0.37 (95% CI: 0.20–0.67) for COVID-19 hospitalization, 0.42 (0.34–0.53) for ED/UC visits, and 0.42 (0.27–0.66) for outpatient visits. Compared to the unvaccinated, those who had received only older versions of COVID-19 vaccines did not show significantly reduced risk of COVID-19 outcomes, including hospital admission.

**Conclusions and Relevance:** Our findings reaffirm current recommendations for broad age-based use of annually updated COVID-19 vaccines given that (1) XBB1.5-adapted vaccines provided significant additional protection against a range of COVID-19 outcomes and (2) older versions of COVID-19 vaccines offered little, if any, additional protection, including against hospital admission, regardless of the number or type of prior doses received.

**KEY POINTS:** 

**Questions:** Does receiving the BNT162b2 XBB1.5-adapted vaccine offer additional protection against COVID-19 hospital admission and ambulatory visits in US adults ≥18 years of age compared to not receiving an XBB1.5-adapted vaccine of any kind? Do older versions of COVID-19 vaccine still provide any protection compared to the unvaccinated?

**Findings:** The BNT162b2 XBB1.5-adapted vaccine (Pfizer-BioNTech 2023–2024 formulation) provided significant additional protection against a range of COVID-19 outcomes during a period when XBB sub-lineages were predominant but JN.1 was also co-circulating and rapidly increasing in prevalence. Older versions of COVID-19 vaccines offered little, if any, additional protection compared to the unvaccinated, including against COVID-19 hospital admission, regardless of the number or type of prior doses received.

**Meaning:** Our findings reaffirm current recommendations for broad age-based use of annually updated COVID-19 vaccines.

## Introduction

XBB and its sub-lineages have been the predominant circulating SARS-CoV-2 strains in the United States since January 2023.^1^ XBB sub-lineages are antigenically and phylogenetically distinct from Omicron BA4 and BA5 sub-lineages that were predominant in late 2022.^1^ Thus, on September 11, 2023, the US Food and Drug Administration authorized or approved updated monovalent mRNA COVID-19 vaccines targeting the XBB1.5 sub-lineage for individuals aged 6 months through 11 years of age and 12 years and older, respectively.^2^ This included use of XBB1.5-adapted vaccines for all recommended doses, including primary series and booster doses.^2^ XBB1.5-adapted vaccines were made broadly available in the United States on September 15, 2023, following Centers for Disease Control and Prevention (CDC) recommendations for use in all individuals aged 6 months and older in preparation for the winter respiratory virus season.^3^ Studies evaluating the association between receipt of XBB1.5-adapted vaccines and the development of clinically relevant COVID-19 endpoints are needed.

## Methods

### Design, Setting, and Participants

Similar to our previous reports,^4–8^ we performed a test-negative case-control study to compare the odds of receipt of a Pfizer-BioNTech BNT162b2 XBB1.5-adapted monovalent vaccine (2023–2024 formulation) between COVID-19 cases and test-negative controls. We included patients ≥18 years of age at Kaiser Permanente Southern California (KPSC) who were diagnosed with acute respiratory infection (ARI; **Table S1**) and tested for SARS-CoV-2 using polymerase chain reaction (PCR) during a hospital admission or emergency department (ED), urgent care (UC), or in-person outpatient encounter from October 10, 2023 through December 10, 2023. The study start date corresponded to 14 days after the date that XBB1.5-adapted vaccines were made available in KPSC (i.e., September 25, 2023). During the study period, XBB sub-lineages were predominantly circulating. JN.1, however, which is a BA2.86 sub-lineage, also began co-circulating and rapidly increasing in prevalence after mid-November.^1^

Participants were required to have ≥1 year of health plan membership (allowing for a 31-day gap in membership to account for delays in membership renewal) to determine comorbidities and medical history. Encounters in which the patient had the following were excluded: (1) another positive SARS-CoV-2 test ≤90 days ago, (2) received any type of XBB1.5-adapted vaccine other than BNT162b2 XBB1.5-adapted vaccine, (3) received a XBB1.5-adapted vaccine ≤2 months after a prior COVID-19 dose, (4) received a BNT162b2 XBB1.5-adapted vaccine ≤14 days prior to the encounter, (5) received any other non-XBB1.5-adapted booster doses (e.g., BA.4/5- adapted bivalent or wild-type boosters) outside of CDC recommended dosing intervals (i.e., defined as receipt of any mRNA BA4.5 bivalent dose between August 31, 2022 and September 11, 2023 with ≥8 weeks [≥56 days] since their most recent dose of original wild-type COVID-19 mRNA vaccine received and a minimal required interval of ≥28 days between a second and subsequent wild-type dose), (6) received nirmatrelvir/ritonavir or any other COVID-19 outpatient antiviral or monoclonal antibody (i.e., molnupiravir, remdesivir, bebtelovimab, bamlanivimab, casirivimab, cilgavimab, sotrovimab, tixagevimab) in the 30 days prior to a COVID-19 encounter, or (7) a hospital admission that, despite having an ARI diagnosis with a positive SARS-CoV-2 test, was determined to be likely unrelated to COVID-19 or clearly related to another cause based on medical chart review that was conducted by trained research staff who were blinded to vaccination status and later validated by a blinded physician investigator (BKA; **Supplemental Appendix**).

### Outcomes

Cases were those with a positive SARS-CoV-2 PCR test, and controls tested negative. SARS-CoV-2 PCR tests among cases and controls were restricted to those administered ≤14 days prior to the initial ARI encounter through ≤3 days after the encounter. Patients could contribute ≥1 event to the study if events were >30 days apart.

### Exposures

All members were eligible for COVID-19 vaccines at no cost based on FDA authorized or approved indications. KPSC EHRs captured all vaccinations administered within the health system. Records were supplemented with vaccine administration data from the California Immunization Registry, to which all healthcare providers are required by law to report COVID- 19 vaccinations within 24 hours. As such, misclassification of vaccination status was unlikely. To be considered vaccinated, the dose had to occur >14 days before testing for SARS-CoV-2. For the primary analysis, COVID-19 outcomes were evaluated comparing individuals who received BNT162b2 XBB1.5-adapted vaccine to those who did not receive an XBB1.5-adapted vaccine of any kind (including the unvaccinated), regardless of prior COVID-19 vaccination or SARS-CoV-2 infection history. Additional analyses compared those who had received BNT162b2 XBB1.5-adapted vaccine to those who had received (1) ≥1 dose of BA.4/5-adapted bivalent vaccine but no XBB1.5-adapted vaccine of any kind, (2) ≥3 or ≥2 doses of original wild-type mRNA vaccine but no variant-adapted vaccines of any kind (e.g., XBB1.5-adapted or BA.4/5-adapted bivalent doses), and (3) the unvaccinated. Among those who did not receive an XBB1.5-adapted vaccine of any kind, we also evaluated the same COVID-19 outcomes comparing individuals who received ≥1 dose of BA.4/5-adapted bivalent vaccine or who received ≥3 or ≥2 original wild-type COVID-19 vaccine doses without a bivalent vaccine of any kind to the unvaccinated. These comparisons were used to estimate remaining protection from prior non-XBB1.5-adapted vaccines (i.e., either BA.4/5-adapted or wild-type doses) during the study period.

### Statistical Analysis

Odds ratios (ORs) and 95% confidence intervals (CIs; based on the Wald method) were calculated from multivariable logistic regression models that included week of encounter, age (18−49, 50−64, and ≥65 years), sex (female, male), self-reported race/ethnicity (non-Hispanic Asian/Pacific Islander, non-Hispanic African-American/Black, Hispanic/Latinx, non-Hispanic White, other [including individuals who identified as American Indian or multiple or other race and ethnicity], and unknown), body mass index (<18.5, 18.5−24.9, 25.0−29.9, 30.0−34.9, ≥35.0 kg/m², and unknown), Charlson risk score (0, 1, 2, 3, and ≥4), receipt of influenza vaccine in the year before admission (yes or no), receipt of pneumococcal vaccine in the 5 years before admission (yes or no), health-care utilization in the year before admission (i.e., number of hospital admissions and ED or outpatient visits), and documentation of previous SARS-CoV-2 infection confirmed by PCR or antigen test (ever *vs* never) for pre-delta, delta, and omicron periods. Missing values were treated as separate categories for all variables in all analyses.

Analyses were done separately for hospital admissions, ED/UC encounters, and outpatient visits. Primary analyses were further stratified by age group (18–64 *vs* ≥65 years). In sensitivity analyses, we examined the impact on the primary analysis of including patients who received COVID-19 antiviral or monoclonal antibody treatment. Analyses were performed using SAS version 9.4. This study was approved by the KPSC institutional review board which waived the requirement for informed consent.

## Results

Of 26,187 ARI encounters among adults ≥18 years of age with continuous enrollment and an eligible SARS-CoV-2 PCR test, 24,007 met study selection criteria (**Figure 1**; 2980 [12.4%] hospital admissions, 15,255 [63.5%] ED/UC, and 5624 [23.4%] outpatient visits). Median age was 54 (interquartile range: 37–70). Overall, 17.6% (4232 / 24,007) tested positive for SARS-CoV-2 and 6.6% (1583 / 24,007) received BNT162b2 XBB1.5-adapted vaccine. Of 4232 cases and 19,775 controls, 171 (4.1%) and 1412 (7.4%), respectively, received BNT162b2 XBB1.5- adapted vaccine (**Table 1**). 22,424 (93.4%) never received an XBB1.5-adapted COVID-19 vaccine of any kind and 2717 (11.3%) never received a COVID-19 vaccine of any kind. Among those who received BNT162b2 XBB1.5-adapted vaccine, median time since receipt of their most recent previous dose of COVID-19 vaccine was 363 days (range: 63–998). Overall, median time since receipt of an XBB1.5-adapted vaccine was 30 days (range: 14–73). Adjusted ORs for testing positive for SARS-CoV-2 among those who received a BNT162b2 XBB1.5-adapted vaccine (*vs* not having received an XBB1.5-adapted vaccine of any kind) were 0.37 (95% CI: 0.20–0.67) for COVID-19 hospital admission, 0.42 (0.34–0.53) for ED/UC visits, and 0.42 (0.27–0.66) for outpatient visits (**Figure 2, Table S2**). In stratified analyses, reductions in risk of testing positive for SARS-CoV-2 among those who received a BNT162b2 XBB1.5-adapted vaccine were similar regardless of comparison group, including those who received (1) ≥1 BA.4/5-adapted bivalent vaccine and no XBB1.5-adapted vaccine, (2) ≥3 or ≥2 doses of original wild-type vaccine without any variant-adapted boosters of any kind (e.g., BA.1-adapted or BA.4/5-adapted bivalent vaccines or XBB1.5-adapted vaccines), and (3) the unvaccinated, across all settings of care (i.e., hospital admission and ED/UC or outpatient visits; **Figure 2, Table S2**). Overall, results also appeared generally similar across age groups 18–64 and ≥65 years with a trend toward greater risk reduction for age ≥65 for outpatient encounters; however, CIs were wide for 18–64-year-olds which made age-group comparisons difficult (**Tables S2, S3**).

**Figure 1.**
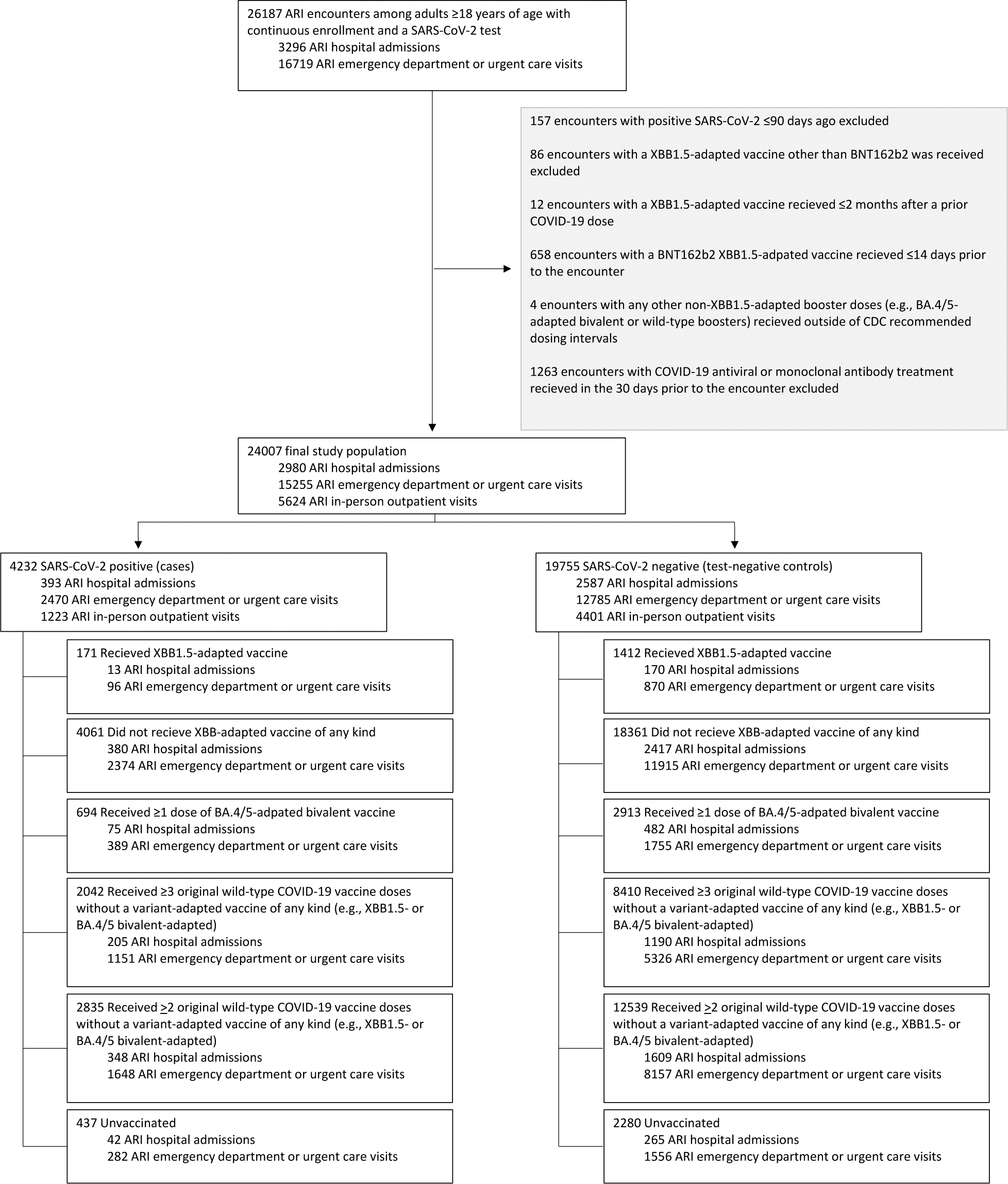
Selection criteria ARI = acute respiratory infection. CDC = Centers for Disease Control and Prevention.

**Figure 2.**
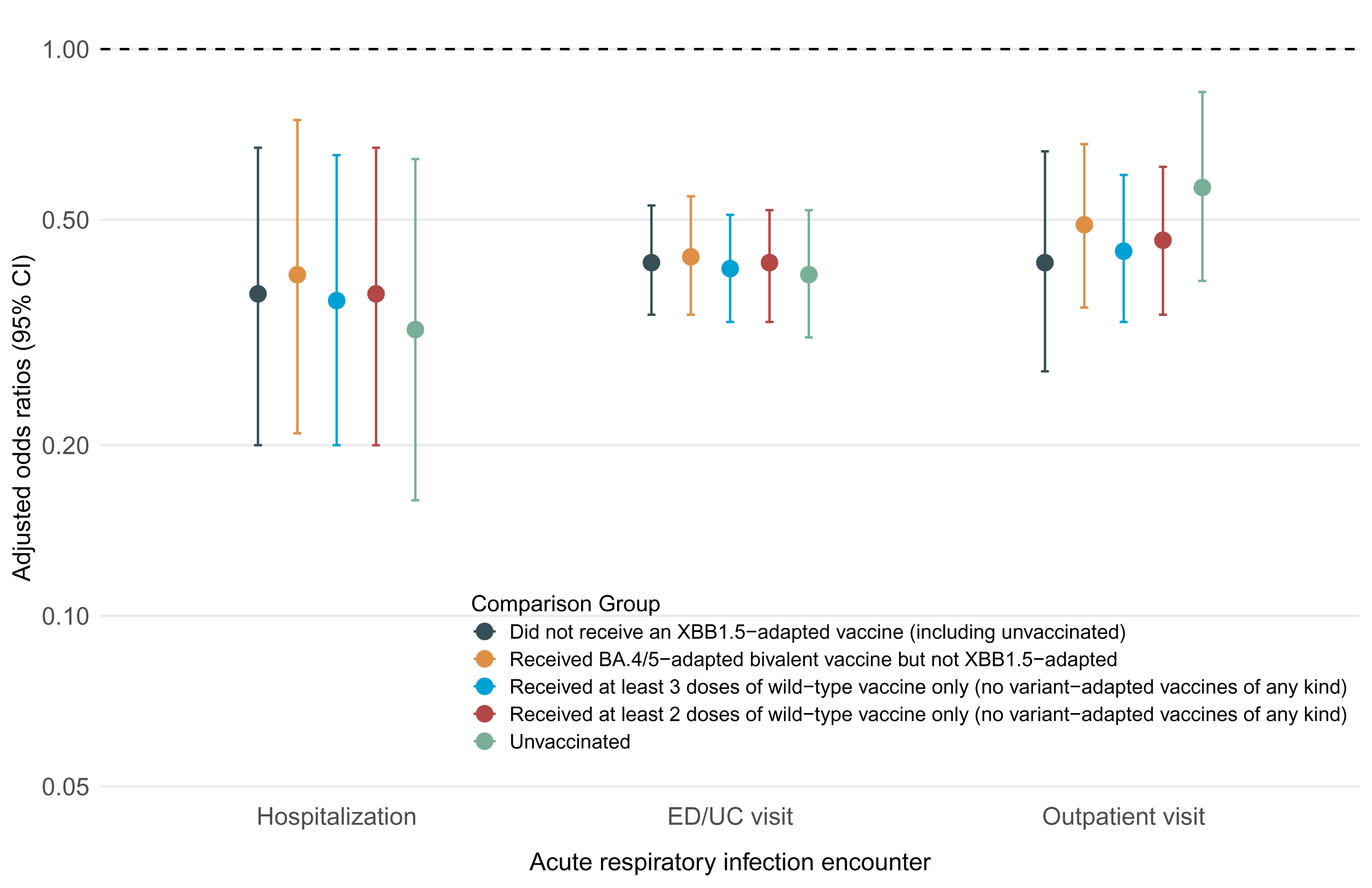
Risk of COVID-19 outcome among those who received BNT162b2 XBB1.5-adapted vaccine by comparison group, adults ≥18 years of age CI = confidence interval. ED = emergency department; UC = urgent care. Models adjusted for week of encounter, age, sex, self-reported race/ethnicity, BMI, Charlson comorbidity index, prior SARS-CoV-2 infection, and utilization history (flu and pneumococcal vaccination, inpatient, ED, and outpatient encounters in prior year).

**Table 1:** Characteristics of Study Population.

| Characteristic | Did not receive an XBB1.5-adapted vaccine of any kind | Received BNT162b2 XBB1.5-adapted vaccine | SARS-CoV-2 Positive | SARS-CoV-2 Negative | Total |
| --- | --- | --- | --- | --- | --- |
| **Total N** | 22424 | 1583 | 4232 | 19775 | 24007 |
| **Prior COVID-19 vaccination** |  |  |  |  |  |
| Unvaccinated | 2717 (12.1%) | 0 (0%) | 437 (10.3%) | 2280 (11.5%) | 2717 (11.3%) |
| 1 original wild-type dose and no BA.4/5-adapted bivalent doses | 713 (3.2%) | 0 (0%) | 95 (2.2%) | 618 (3.1%) | 713 (3.0%) |
| 2 original wild-type doses and no BA.4/5-adapted bivalent doses | 4922 (22.0%) | 32 (2.0%) | 797 (18.8%) | 4157 (21.0%) | 4954 (20.6%) |
| ≥3 original wild-type doses and no BA.4/5-adapted bivalent dose | 10452 (46.6%) | 757 (47.8%) | 2119 (50.1%) | 9090 (46.0%) | 11209 (46.7%) |
| ≥1 BA.4/5-adapted bivalent doses | 3620 (16.1%) | 794 (50.2%) | 784 (18.5%) | 3630 (18.4%) | 4414 (18.4%) |
| **Age in years at time of encounter** |  |  |  |  |  |
| 18–49 | 9962 (44.4%) | 311 (19.6%) | 1562 (36.9%) | 8711 (44.1%) | 10273 (42.8%) |
| 50–64 | 5274 (23.5%) | 294 (18.6%) | 1020 (24.1%) | 4548 (23%) | 5568 (23.2%) |
| ≥65 | 7188 (32.1%) | 978 (61.8%) | 1650 (39%) | 6516 (33%) | 8166 (34%) |
| **Sex** |  |  |  |  |  |
| Female | 13709 (61.1%) | 885 (55.9%) | 2601 (61.5%) | 11993 (60.6%) | 14594 (60.8%) |
| Male | 8715 (38.9%) | 698 (44.1%) | 1631 (38.5%) | 7782 (39.4%) | 9413 (39.2%) |
| **Self-Reported Race / Ethnicity** |  |  |  |  |  |
| Non-Hispanic African American/Black | 2321 (10.4%) | 251 (15.9%) | 466 (11%) | 2106 (10.6%) | 2572 (10.7%) |
| Non-Hispanic Asian/Pacific Islander | 2521 (11.2%) | 150 (9.5%) | 509 (12%) | 2162 (10.9%) | 2671 (11.1%) |
| Hispanic/Latinx | 10340 (46.1%) | 485 (30.6%) | 1883 (44.5%) | 8942 (45.2%) | 10825 (45.1%) |
| Non-Hispanic White | 970 (4.3%) | 57 (3.6%) | 171 (4%) | 856 (4.3%) | 1027 (4.3%) |
| Other/Unknown* | 6272 (28%) | 640 (40.4%) | 1203 (28.4%) | 5709 (28.9%) | 6912 (28.8%) |
| **Encounter type** |  |  |  |  |  |
| Hospital admission | 2942 (13.1%) | 186 (11.7%) | 539 (12.7%) | 2589 (13.1%) | 3128 (13%) |
| Emergency department visit | 6444 (28.7%) | 424 (26.8%) | 1286 (30.4%) | 5582 (28.2%) | 6868 (28.6%) |
| Urgent care visit | 7845 (35%) | 542 (34.2%) | 1184 (28%) | 7203 (36.4%) | 8387 (34.9%) |
| In-person outpatient visit | 5193 (23.2%) | 431 (27.2%) | 1223 (28.9%) | 4401 (22.3%) | 5624 (23.4%) |
| **Charlson comorbidity index** |  |  |  |  |  |
| 0 | 10562 (47.1%) | 456 (28.8%) | 1859 (43.9%) | 9159 (46.3%) | 11018 (45.9%) |
| 1 | 4227 (18.9%) | 285 (18%) | 770 (18.2%) | 3742 (18.9%) | 4512 (18.8%) |
| 2 | 2088 (9.3%) | 227 (14.3%) | 445 (10.5%) | 1870 (9.5%) | 2315 (9.6%) |
| 3 | 1333 (5.9%) | 157 (9.9%) | 270 (6.4%) | 1220 (6.2%) | 1490 (6.2%) |
| ≥4 | 4214 (18.8%) | 458 (28.9%) | 888 (21%) | 3784 (19.1%) | 4672 (19.5%) |
| **Body mass index (BMI; kg/m<sup>2</sup>)** |  |  |  |  |  |
| Underweight (<18.5) | 488 (2.2%) | 18 (1.1%) | 84 (2%) | 422 (2.1%) | 506 (2.1%) |
| Normal or healthy weight (18.5–24.9) | 5038 (22.5%) | 432 (27.3%) | 993 (23.5%) | 4477 (22.6%) | 5470 (22.8%) |
| Overweight (25.0–29.9) | 6737 (30%) | 514 (32.5%) | 1349 (31.9%) | 5902 (29.8%) | 7251 (30.2%) |
| Obese, class 1 (30.0–34.9) | 5104 (22.8%) | 363 (22.9%) | 936 (22.1%) | 4531 (22.9%) | 5467 (22.8%) |
| Obese, class 2-3 (≥35.0) | 4830 (21.5%) | 251 (15.9%) | 835 (19.7%) | 4246 (21.5%) | 5081 (21.2%) |
| Unknown | 227 (1%) | 5 (0.3%) | 35 (0.8%) | 197 (1%) | 232 (1%) |
| **Prior documented positive SARS-CoV-2 PCR or antigen tests** |  |  |  |  |  |
| Pre-delta era (March 1, 2020 – June 11, 2021) | 2445 (10.9%) | 88 (5.6%) | 437 (10.3%) | 2096 (10.6%) | 2533 (10.6%) |
| Delta era (June 12, 2021 – December 20, 2021) | 685 (3.1%) | 24 (1.5%) | 103 (2.4%) | 606 (3.1%) | 709 (3%) |
| Omicron era (December 21, 2021 – current) | 6458 (28.8%) | 412 (26%) | 976 (23.1%) | 5894 (29.8%) | 6870 (28.6%) |
| **Healthcare Utilization (counts) in year prior to encounter** |  |  |  |  |  |
| Outpatient Visits |  |  |  |  |  |
| 0 | 1216 (5.4%) | 3 (0.2%) | 174 (4.1%) | 1045 (5.3%) | 1219 (5.1%) |
| 1 | 1291 (5.8%) | 31 (2%) | 220 (5.2%) | 1102 (5.6%) | 1322 (5.5%) |
| 2–4 | 4349 (19.4%) | 138 (8.7%) | 748 (17.7%) | 3739 (18.9%) | 4487 (18.7%) |
| 5–9 | 5616 (25%) | 358 (22.6%) | 1018 (24.1%) | 4956 (25.1%) | 5974 (24.9%) |
| ≥10 | 9952 (44.4%) | 1053 (66.5%) | 2072 (49%) | 8933 (45.2%) | 11005 (45.8%) |
| Emergency Department Visits |  |  |  |  |  |
| 0 | 14255 (63.6%) | 1018 (64.3%) | 2729 (64.5%) | 12544 (63.4%) | 15273 (63.6%) |
| 1 | 4173 (18.6%) | 290 (18.3%) | 789 (18.6%) | 3674 (18.6%) | 4463 (18.6%) |
| ≥2 | 3996 (17.8%) | 275 (17.4%) | 714 (16.9%) | 3557 (18%) | 4271 (17.8%) |
| Hospital Admissions |  |  |  |  |  |
| 0 | 19405 (86.5%) | 1351 (85.3%) | 3701 (87.5%) | 17055 (86.2%) | 20756 (86.5%) |
| 1 | 1895 (8.5%) | 163 (10.3%) | 339 (8%) | 1719 (8.7%) | 2058 (8.6%) |
| ≥2 | 1124 (5%) | 69 (4.4%) | 192 (4.5%) | 1001 (5.1%) | 1193 (5%) |
| **Received influenza vaccine in year prior to encounter** |  |  |  |  |  |
| No | 11462 (51.1%) | 81 (5.1%) | 1888 (44.6%) | 9655 (48.8%) | 11543 (48.1%) |
| Yes | 10962 (48.9%) | 1502 (94.9%) | 2344 (55.4%) | 10120 (51.2%) | 12464 (51.9%) |
| **Received pneumococcal vaccine in 5 years prior to encounter** |  |  |  |  |  |
| No | 17107 (76.3%) | 1016 (64.2%) | 3175 (75%) | 14948 (75.6%) | 18123 (75.5%) |
| Yes | 5317 (23.7%) | 567 (35.8%) | 1057 (25%) | 4827 (24.4%) | 5884 (24.5%) |

\* Race/ethnicity is reported by participant. “Other” includes individuals who self-identified as American Indian or “multiple” or “other” race and ethnicity. Race/ethnicity was included as a confounder due to associations with vaccination and COVID-19 healthcare seeking behavior and outcomes.

In sensitivity analyses, results were similar when patients who received antiviral or monoclonal antibody treatment in the 30 days prior to their COVID-19 encounter (n=1263) were included (**Table S4)**. The most common reasons for hospital admission in cases that were excluded based on chart review (i.e., likely unrelated to COVID-19; n=198) were labor and delivery (13/198 [6.6%]), injuries requiring urgent medical care (10/198 [5.1%]), surgery/neurosurgery (32/198 [16.2%]), sepsis due to another etiology (e.g., urinary tract infection, bacteremia, or other infection; 41/198 [20.7%]), and medical conditions unrelated to possible COVID-19-associated conditions (e.g., bacterial infection without sepsis, malignancy, drug withdrawal, seizures: 93/198 [47.0%]).

Finally, compared to the unvaccinated, individuals who had not received an XBB1.5-adapted vaccine of any kind but had received older versions of COVID-19 vaccines (i.e., ≥1 BA.4/5- adapted bivalent dose or ≥3 or ≥2 original wild-type doses and no variant-adapted vaccines of any kind) did not show significantly reduced risk of COVID-19 outcomes, including hospital admission, during the study period (**Figure 3**).

**Figure 3.**
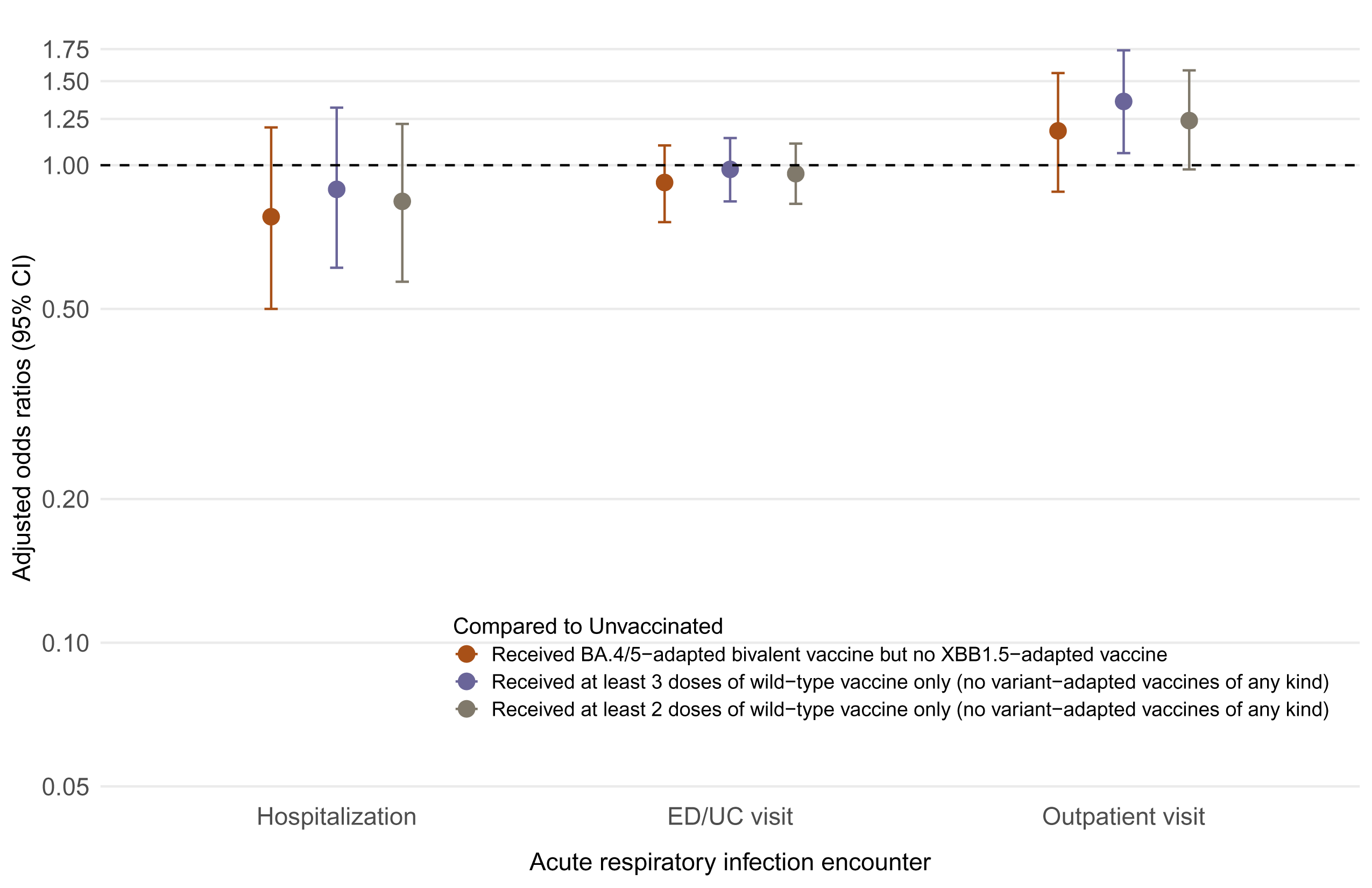
Risk of COVID-19 outcome among those who received only older (non-XBB1.5- adapted) versions of COVID-19 vaccines compared to the unvaccinated by vaccine regimen received, adults ≥18 years of age CI = confidence interval. ED = emergency department; UC = urgent care. Models adjusted for week of encounter, age, sex, self-reported race/ethnicity, BMI, Charlson comorbidity index, prior SARS-CoV-2 infection, and utilization history (flu and pneumococcal vaccination, inpatient, ED, and outpatient encounters in prior year).

## Discussion

In this test-negative case-control study conducted in a large US healthcare system during the early part of the 2023–2024 viral respiratory season in the United States, compared to individuals who did not receive a XBB1.5-adapted vaccine of any kind, those who received a BNT162b2 XBB1.5-adapted vaccine had a 63% (95%CI: 33–80%) reduced risk of COVID-19 hospital admission, 58% (47–66%) lower risk of COVID-19 ED/UC encounters, and 58% (34–73%) lower risk of in-person outpatient visits for COVID-19 after a median of 30 days since receipt of the XBB1.5-adapted dose. XBB sub-lineages were predominant during our study period, however, during the last several weeks of our study when many cases accrued, JN.1 was rapidly increasing in prevalence across the United States and in the State of California.^1^ Preliminary data suggest that BA2.86 and JN.1 show signs of slightly increased immune evasion to XBB1.5- adapted vaccines, however, both strains are still neutralized by BNT162b2 XBB1.5-adapted vaccine.^9,10^ Our results help confirm these early pre-clinical studies^9,10^ by showing that XBB1.5- adapted vaccine provided additional protection against COVID-19 during a period of XBB and JN.1 co-circulation. However, the majority of cases in our analyses were likely XBB sub- lineages.

Consistent with previous reports of COVID-19 vaccine effectiveness,^4–8^ estimated reductions in risk appeared higher for the more severe outcome of hospital admission across all comparison groups (OR point estimates 0.32–0.37) as compared to less severe outcomes like ED/UC and outpatient visits (ORs 0.40–0.57), although CIs overlapped. Importantly, BNT162b2 XBB1.5- adapted vaccine provided similar additional protection in adults regardless of age group and the number of prior COVID-19 vaccine doses received for all COVID-19 outcomes. This latter finding was consistent with our results suggesting that prior receipt of only older versions of COVID-19 vaccines (i.e., receipt of BA.4/5-adapted bivalent vaccine or ≥3 or ≥2 original wild-type doses but no XBB1.5-adapted vaccine) provided little, if any, current additional protection compared to the unvaccinated against COVID-19 outcomes, including hospital admission. Thus, analogous to influenza, although older versions of COVID-19 vaccines once provided high levels of protection, the combination of waning vaccine-induced immunity and continuous SARS-CoV-2 strain evolution eventually renders prior versions of vaccines ineffective. This, in turn, warrants routine updates to COVID-19 vaccines—also like influenza—so long as SARS-CoV-2 continues to circulate and cause disease. For the week ending December 9, 2023, there were 23,432 new COVID-19 hospital admissions in the United States—the highest rate since last winter.^11^ This still represents a large public health burden and is roughly three times higher than the number of new weekly influenza hospitalizations that occurred during the same period (7090);^12^ albeit fewer than the approximately 150,000 weekly hospitalizations seen at the peak of the Omicron BA.1 wave.^11^ Our findings help reaffirm current recommendations for broad age-based use of annually updated COVID-19 vaccines in the United States prior to likely winter peaks in disease activity.^13^

Uptake of XBB1.5-adapted vaccines in the United States to date has been low. As of December 22, 2023 only 19% and 37% of all adults ≥18 and ≥65 years of age, respectively, had received an XBB1.5-adapted vaccine.^14^ Current COVID-19 vaccine coverage significantly lags that of seasonal influenza vaccines, despite both vaccines being made available during the autumn and winter, and current CDC guidelines that support co-administration of the two vaccines.^15^ Reasons for low COVID-19 vaccine uptake likely include reduced concern about COVID-19 in the general population over time and as pandemic declarations ended, annual COVID-19 vaccination not yet being seen as a “routine health activity,” confusion about risk level regarding COVID-19, and continued skepticism in some populations about the safety and effectiveness of mRNA vaccines.^16^

A recent preprint study showed that among adults ≥65 years of age in Denmark, receipt of an XBB1.5-adapted vaccine (90% of which were the BNT162b2 XBB1.5-adapted vaccine in the study population) led to a 75% (95%CI: 61–84%) reduced risk of COVID-19 hospital admission over an average follow-up time of 10 days compared to those who did not receive an XBB1.5- adapted vaccine^17^—an estimate that was comparable to ours, albeit slightly higher, against the same outcome and comparison group. However, this study had shorter follow-up time and included data only through the end of October, which was prior to the emergence and rapid growth of the JN.1 strain. Another recent preprint study conducted in the Netherlands also recently showed similar reductions in risk of hospital admission (71% [67–74%]) among adults ≥60 years of age who were previously vaccinated.^18^ Our study helps confirm these early global findings but also describes the association between XBB1.5-adapted vaccine receipt and the development of COVID-19 across a broader range of outcomes and age groups, in a more diverse study population, and during a more recent time period that included when the JN.1 strain was rapidly increasing in prevalence in the United States.

Our study has limitations. Although we controlled for key sociodemographic and clinical characteristics, there may be residual confounding driven by unaccounted for differences in the likelihood of exposure or severity of SARS-CoV-2 infection between vaccinated and unvaccinated individuals. Although individuals who are more likely to get vaccinated against COVID-19 may also be more likely to seek care or testing for SARS-CoV-2, the test-negative design of our study helps mitigate against bias caused by differences in healthcare-seeking behavior—including the propensity to test.^19–21^ We also controlled for prior healthcare utilization, age, and underlying comorbid illness to help mitigate the potential for healthy vaccinee bias. A second limitation is that median time since receipt of an XBB1.5-adapted vaccine was only 30 days, and future studies are needed to evaluate durability of protection. In addition, although we know our study was conducted during a period when XBB sub-lineages were predominant, but JN.1 was also co-circulating and rapidly increasing in prevalence, we did not have genotype information available for all our cases, nor were we able to estimate sub- lineage-specific estimates. Thus, future studies describing the association between receipt of XBB1.5-adapted vaccines and development of strain-specific BA2.86 sub-lineage-related disease (e.g., JN.1) are needed. Another limitation is that there was likely misclassification of previous infections, particularly among those who did not seek testing at KPSC. If undocumented previous infection was more likely in unvaccinated individuals, for example, this could contribute to underestimation of vaccine protection. It remains possible that some healthcare encounters were “with COVID-19” rather than “for COVID-19,” and this could lead to underestimation of the protective effects of vaccination against medically attended disease. To help mitigate this bias, we (1) used medical record review to exclude hospital admissions that were unrelated to COVID-19, and (2) restricted our analyses to patients presenting with ARI for all outcomes.

In conclusion, individuals who did not receive an XBB1.5-adapted vaccine and had received only older versions of COVID-19 vaccines had little, if any, additional protection compared to the unvaccinated against COVID-19 endpoints, including hospital admission, regardless of the number or type of prior doses received. Receipt of a BNT162b2 XBB1.5-adapted vaccine, however, was associated with significantly reduced risk of developing a range of COVID-19 outcomes during the early part of the 2023–2024 viral respiratory season—with the strongest protective effects seen against hospital admission. These two findings help reaffirm current recommendations for broad age-based use of annually updated COVID-19 vaccines. Uptake of this 2023–2024 formulation of COVID-19 vaccines, however, remains low, and targeted and tailored interventions to continuously improve annual COVID-19 uptake are warranted.

### Data statement

Anonymized data that support the findings of this study may be made available from the investigative team in the following conditions: (1) agreement to collaborate with the study team on all publications, (2) provision of external funding for administrative and investigator time necessary for this collaboration, (3) demonstration that the external investigative team is qualified and has documented evidence of training for human subjects protections, and (4) agreement to abide by the terms outlined in data use agreements between institutions.

### Role of the Funding Source

This study was sponsored by Pfizer. The study design was jointly developed by KPSC and Pfizer. KPSC collected and analyzed the data. Pfizer did not participate in the collection or analysis of data. KPSC and Pfizer participated in the interpretation of data, in the writing of the report, and in the decision to submit the paper for publication.

## Acknowledgements

TBF and JMS had full access to all the data in the study and takes responsibility for the integrity of the data and the accuracy of the data analysis.

The authors would like to acknowledge Fagen Xie, PhD and Harpreet Takhar, MPH from KPSC Department of Research & Evaluation. We also thank Joann M. Zamparo, MPH who is an employee of Pfizer Inc and did not receive compensation beyond her salary, for her administrative support of this project.

LJ, LP, and JMM are employees of and hold stock and/or stock options in Pfizer Inc. SYT, TF, JMS, VH, SS, JAS and BA received research support from Pfizer during the conduct of this study that was paid directly to KPSC. BA received research support for work unrelated to this study provided by Pfizer, Moderna, Dynavax, Seqirus, GlaxoSmithKline and Genentech. JMS received research support from ALK, Inc., Dynavax, and Novavax for work unrelated to this study. TBF previously owned stock in Pfizer Inc. SYT received research support from GSK and Genentech for work unrelated to this study.

## Notes

### Author Declarations

This study was approved by the Kaiser Permanente Southern California Institutional Review Board which waived the requirement for informed consent.

## References

1. 1. U.S. Centers for Disease Control and Prevention. COVID Data Tracker: Monitoring Variant Proportions. Accessed December 18, 2023, https://covid.cdc.gov/covid-data-tracker/#variant-proportions

2. U.S. Food and Drug Administration. COVID-19 Vaccines for 2023-2024. Accessed December 18, 2023, https://www.fda.gov/emergency-preparedness-and-response/coronavirus-disease-2019-covid-19/covid-19-vaccines-2023-2024

3. Regan JJ, Moulia DL, Link-Gelles R, et al. Use of Updated COVID-19 Vaccines 2023-2024 Formula for Persons Aged >/=6 Months: Recommendations of the Advisory Committee on Immunization Practices - United States, September 2023. MMWR Morb Mortal Wkly Rep. Oct 20 2023;72(42):1140- 1146. doi:10.15585/mmwr.mm7242e1

4. Tartof SY, Frankland TB, Slezak JM, et al. Effectiveness Associated With BNT162b2 Vaccine Against Emergency Department and Urgent Care Encounters for Delta and Omicron SARS-CoV-2 Infection Among Adolescents Aged 12 to 17 Years. JAMA Netw Open. Aug 1 2022;5(8):e2225162. doi:10.1001/jamanetworkopen.2022.25162

5. Tartof SY, Frankland TB, Slezak JM, Puzniak L, Ackerson BK, Jodar L, McLaughlin JM. Receipt of BNT162b2 Vaccine and COVID-19 Ambulatory Visits in US Children Younger Than 5 Years. JAMA. Oct 3 2023;330(13):1282–1284. doi:10.1001/jama.2023.17473

6. Tartof SY, Slezak JM, Puzniak L, et al. BNT162b2 vaccine effectiveness against SARS-CoV-2 omicron BA.4 and BA.5. Lancet Infect Dis. Dec 2022;22(12):1663-1665. doi:10.1016/S1473-3099(22)00692-2

7. Tartof SY, Slezak JM, Puzniak L, et al. Effectiveness of BNT162b2 BA.4/5 bivalent mRNA vaccine against a range of COVID-19 outcomes in a large health system in the USA: a test-negative case-control study. Lancet Respir Med. Dec 2023;11(12):1089-1100. doi:10.1016/S2213-2600(23)00306-5

8. Tartof SY, Slezak JM, Puzniak L, et al. Effectiveness and durability of BNT162b2 vaccine against hospital and emergency department admissions due to SARS-CoV-2 omicron sub-lineages BA.1 and BA.2 in a large health system in the USA: a test-negative, case-control study. Lancet Respir Med. Feb 2023;11(2):176-187. doi:10.1016/S2213-2600(22)00354-X

9. Modjarrad K, Che Y, Chen W, et al. Preclinical Characterization of the Omicron XBB.1.5-Adapted BNT162b2 COVID-19 Vaccine. bioRxiv. 2023:2023.11.17.567633. doi:10.1101/2023.11.17.567633

10. Wang Q, Guo Y, Bowen A, et al. XBB.1.5 monovalent mRNA vaccine booster elicits robust neutralizing antibodies against emerging SARS-CoV-2 variants. bioRxiv. 2023:2023.11.26.568730. doi:10.1101/2023.11.26.568730

11. U.S. Centers for Disease Control and Prevention. United States COVID-19 Hospitalizations, Deaths, Emergency Department (ED) Visits, and Test Positivity by Geographic Area. Accessed December 18, 2023, https://covid.cdc.gov/covid-data-tracker/#maps_new-admissions-rate-county

12. U.S. Centers for Disease Control and Prevention. Weekly U.S. Influenza Surveillance Report: New Hospital Admissions Reported to National Healthcare Safety Network (NHSN), Updated December 15, 2023. Accessed December 18, 2023, https://www.cdc.gov/flu/weekly/index.htm#:~:text=CDC%20estimates%20that%20there%20have,there%20are%20still%20vaccines%20available.

13. Wiemken TL, Khan F, Puzniak L, et al. Seasonal trends in COVID-19 cases, hospitalizations, and mortality in the United States and Europe. Sci Rep. Mar 8 2023;13(1):3886. doi:10.1038/s41598-023-31057-1

14. U.S. Centers for Disease Control and Prevention. Vaccination Trends—Adults. Accessed December 22, 2023, https://www.cdc.gov/respiratory-viruses/data-research/dashboard/vaccination-trends-adults.html#:~:text=The%20percent%20of%20the%20population%20reporting%20receipt%20of%20the%20updated,)%20among%20adults%20age%2065%2B.

15. U.S. Centers for Disease Control and Prevention. Getting a Flu Vaccine and other Recommended Vaccines at the Same Time. Accessed December 18, 2023, https://www.cdc.gov/flu/prevent/coadministration.htm

16. Sparks G, Kirzinger A, Kearney A, Valdes I. KFF COVID-19 Vaccine Monitor November 2023: With COVID Concerns Lagging, Most People Have Not Gotten Latest Vaccine And Half Say They Are Not Taking Precautions This Holiday Season. November 17, 2023.

17. Hansen CH, Mousten-Helms IR, Rasmussen M, Bolette S, Valentiner-Branth P, Ullum H. Effectiveness of the XBB.1.5 updated COVID-19 vaccine against hospitalisation: a nation-wide cohort study in Denmark, October 2023 (November 8, 2023). *Available at SSRN:* https://ssrncom/abstract=4627268 *or* http://dxdoiorg/102139/ssrn4627268. November 8, 2023;

18. Werkhoven Hv, Valk A-W, Smagge B, et al. Early COVID-19 vaccine effectiveness of XBB.1.5 vaccine against hospitalization and ICU admission, the Netherlands, 9 October - 5 December 2023. *medRxiv*. 2023:2023.12.12.23299855. doi:10.1101/2023.12.12.23299855

19. Haber M, An Q, Foppa IM, Shay DK, Ferdinands JM, Orenstein WA. A probability model for evaluating the bias and precision of influenza vaccine effectiveness estimates from case-control studies. Epidemiol Infect. May 2015;143(7):1417–26. doi:10.1017/S0950268814002179

20. Jackson ML, Nelson JC. The test-negative design for estimating influenza vaccine effectiveness. Vaccine. Apr 19 2013;31(17):2165–8. doi:10.1016/j.vaccine.2013.02.053

21. Lipsitch M, Jha A, Simonsen L. Observational studies and the difficult quest for causality: lessons from vaccine effectiveness and impact studies. Int J Epidemiol. Dec 1 2016;45(6):2060–2074. doi:10.1093/ije/dyw124

