## Supplemental Tables for "BNT162b2 XBB1.5-adapted Vaccine and COVID-19 Hospital Admissions and Ambulatory Visits in US Adults"

### Supplement

**Table S1.** Acute Respiratory Infection Codes, October 10, 2023 through December 10, 2023

| | |
|---|---|
| A48.1 | LEGIONNAIRES' DISEASE |
| B34.2 | CORONAVIRUS INFECTION, UNSPECIFIED |
| B44.0 | INVASIVE PULMONARY ASPERGILLOSIS |
| B97.29 | OTH CORONAVIRUS AS THE CAUSE OF DISEASES CLASSD ELSWHR |
| J00 | ACUTE NASOPHARYNGITIS (COMMON COLD) |
| J01.00 | ACUTE MAXILLARY SINUSITIS, UNSPECIFIED |
| J01.10 | ACUTE FRONTAL SINUSITIS, UNSPECIFIED |
| J01.20 | ACUTE ETHMOIDAL SINUSITIS, UNSPECIFIED |
| J01.30 | ACUTE SPHENOIDAL SINUSITIS, UNSPECIFIED |
| J01.40 | ACUTE PANSINUSITIS, UNSPECIFIED |
| J01.80 | OTHER ACUTE SINUSITIS |
| J01.90 | ACUTE SINUSITIS, UNSPECIFIED |
| J02.0 | STREPTOCOCCAL PHARYNGITIS |
| J02.8 | ACUTE PHARYNGITIS DUE TO OTHER SPECIFIED ORGANISMS |
| J02.9 | ACUTE PHARYNGITIS, UNSPECIFIED |
| J03.00 | ACUTE STREPTOCOCCAL TONSILLITIS, UNSPECIFIED |
| J03.90 | ACUTE TONSILLITIS, UNSPECIFIED |
| J04.0 | ACUTE LARYNGITIS |
| J04.10 | ACUTE TRACHEITIS WITHOUT OBSTRUCTION |
| J05.0 | ACUTE OBSTRUCTIVE LARYNGITIS (CROUP) |
| J05.10 | ACUTE EPIGLOTTITIS WITHOUT OBSTRUCTION |
| J06.0 | ACUTE LARYNGOPHARYNGITIS |
| J06.9 | ACUTE UPPER RESPIRATORY INFECTION, UNSPECIFIED |
| J09.X1 | INFLUENZA DUE TO IDENT NOVEL INFLUENZA A VIRUS W PNEUMONIA |
| J09.X2 | FLU DUE TO IDENT NOVEL INFLUENZA A VIRUS W OTH RESP MANIFEST |
| J10.00 | FLU DUE TO OTH IDENT FLU VIRUS W UNSP TYPE OF PNEUMONIA |
| J10.01 | FLU DUE TO OTH IDENT FLU VIRUS W SAME OTH IDENT FLU VIRUS PN |
| J10.08 | INFLUENZA DUE TO OTH IDENT INFLUENZA VIRUS W OTH PNEUMONIA |
| J10.1 | FLU DUE TO OTH IDENT INFLUENZA VIRUS W OTH RESP MANIFEST |
| J10.2 | INFLUENZA DUE TO OTH IDENT INFLUENZA VIRUS W GI MANIFEST |
| J11.00 | FLU DUE TO UNIDENTIFIED FLU VIRUS W UNSP TYPE OF PNEUMONIA |
| J11.08 | FLU DUE TO UNIDENTIFIED FLU VIRUS W SPECIFIED PNEUMONIA |
| J11.1 | FLU DUE TO UNIDENTIFIED INFLUENZA VIRUS W OTH RESP MANIFEST |
| J12.1 | RESPIRATORY SYNCYTIAL VIRUS PNEUMONIA |
| J12.2 | PARAINFLUENZA VIRUS PNEUMONIA |
| J12.3 | HUMAN METAPNEUMOVIRUS PNEUMONIA |
| J12.81 | PNEUMONIA DUE TO SARS-ASSOCIATED CORONAVIRUS |

| | |
|---|---|
| J12.82 | PNEUMONIA DUE TO CORONAVIRUS DISEASE 2019 |
| J12.89 | OTHER VIRAL PNEUMONIA |
| J12.9 | VIRAL PNEUMONIA, UNSPECIFIED |
| J13 | PNEUMONIA DUE TO STREPTOCOCCUS PNEUMONIAE |
| J14 | PNEUMONIA DUE TO HEMOPHILUS INFLUENZAE |
| J15.0 | PNEUMONIA DUE TO KLEBSIELLA PNEUMONIAE |
| J15.1 | PNEUMONIA DUE TO PSEUDOMONAS |
| J15.20 | PNEUMONIA DUE TO STAPHYLOCOCCUS, UNSPECIFIED |
| J15.211 | PNEUMONIA DUE TO METHICILLIN SUSCEP STAPH |
| J15.212 | PNEUMONIA DUE TO METHICILLIN RESISTANT STAPHYLOCOCCUS AUREUS |
| J15.4 | PNEUMONIA DUE TO OTHER STREPTOCOCCI |
| J15.5 | PNEUMONIA DUE TO ESCHERICHIA COLI |
| J15.6 | PNEUMONIA DUE TO OTHER AEROBIC GRAM-NEGATIVE BACTERIA |
| J15.7 | PNEUMONIA DUE TO MYCOPLASMA PNEUMONIAE |
| J15.8 | PNEUMONIA DUE TO OTHER SPECIFIED BACTERIA |
| J15.9 | UNSPECIFIED BACTERIAL PNEUMONIA |
| J16.8 | PNEUMONIA DUE TO OTHER SPECIFIED INFECTIOUS ORGANISMS |
| J18.0 | BRONCHOPNEUMONIA, UNSPECIFIED ORGANISM |
| J18.1 | LOBAR PNEUMONIA, UNSPECIFIED ORGANISM |
| J18.8 | OTHER PNEUMONIA, UNSPECIFIED ORGANISM |
| J18.9 | PNEUMONIA, UNSPECIFIED ORGANISM |
| J20.2 | ACUTE BRONCHITIS DUE TO STREPTOCOCCUS |
| J20.5 | ACUTE BRONCHITIS DUE TO RESPIRATORY SYNCYTIAL VIRUS |
| J20.6 | ACUTE BRONCHITIS DUE TO RHINOVIRUS |
| J20.8 | ACUTE BRONCHITIS DUE TO OTHER SPECIFIED ORGANISMS |
| J20.9 | ACUTE BRONCHITIS, UNSPECIFIED |
| J22 | UNSPECIFIED ACUTE LOWER RESPIRATORY INFECTION |
| J39.0 | RETROPHARYNGEAL AND PARAPHARYNGEAL ABSCESS |
| J39.1 | OTHER ABSCESS OF PHARYNX |
| J39.2 | OTHER DISEASES OF PHARYNX |
| J39.8 | OTHER SPECIFIED DISEASES OF UPPER RESPIRATORY TRACT |
| J80 | ACUTE RESPIRATORY DISTRESS SYNDROME |
| J96.00 | ACUTE RESPIRATORY FAILURE, UNSP W HYPOXIA OR HYPERCAPNIA |
| J96.01 | ACUTE RESPIRATORY FAILURE WITH HYPOXIA |
| J96.02 | ACUTE RESPIRATORY FAILURE WITH HYPERCAPNIA |
| J96.10 | CHRONIC RESPIRATORY FAILURE, UNSP W HYPOXIA OR HYPERCAPNIA |
| J96.11 | CHRONIC RESPIRATORY FAILURE WITH HYPOXIA |
| J96.12 | CHRONIC RESPIRATORY FAILURE WITH HYPERCAPNIA |
| J96.20 | ACUTE AND CHR RESP FAILURE, UNSP W HYPOXIA OR HYPERCAPNIA |
| J96.21 | ACUTE AND CHRONIC RESPIRATORY FAILURE WITH HYPOXIA |
| J96.22 | ACUTE AND CHRONIC RESPIRATORY FAILURE WITH HYPERCAPNIA |

| | |
|---|---|
| J96.90 | RESPIRATORY FAILURE, UNSP, UNSP W HYPOXIA OR HYPERCAPNIA |
| J96.91 | RESPIRATORY FAILURE, UNSPECIFIED WITH HYPOXIA |
| J96.92 | RESPIRATORY FAILURE, UNSPECIFIED WITH HYPERCAPNIA |
| M35.81 | MULTISYSTEM INFLAMATORY SYNDROME |
| M35.89 | OTHER SPECIFIED SYSTEMIC INVOLVMENT OF CONNECTIVE TISSUE |
| R05.1 | ACUTE COUGH |
| R05.3 | CHRONIC COUGH |
| R05.4 | COUGH SYNCOPE |
| R05.8 | OTHER SPECIFIED COUGH |
| R05.9 | COUGH, UNSPECIFIED |
| R09.2 | RESPIRATORY ARREST |
| R50.9 | FEVER, UNSPECIFIED |
| U07.1 | COVID-19 |

### Approach to medical chart review validation of hospital admissions

#### Summary

Chart reviews focused on the elimination of hospitalizations that are clearly not for COVID-19 from analysis (e.g., hospitalizations with incidental SARS-CoV-2 infection that did not contribute to the need for hospitalization and, therefore, do not reflect severe COVID-19 disease). Extensive chart review is not required in most cases.

Most cases are clearly for COVID-19 or for another etiology with only incidental SARS-CoV-2 infection. Therefore, most chart reviews can be limited to:

1.) Discharge summary and/or
2.) ED note, Admission H&P

However, some cases require more nuanced review and are referred to the MD co-investigator for more detailed review.

3.) Discharge summary
4.) ED note, Admission H&P
5.) Progress notes in limited number of cases
6.) Nursing and other notes in rare cases

#### Definitions

1.) **Clearly COVID-19 associated:**
   a. severe respiratory illness [e.g., acute hypoxemic respiratory failure], severe vomiting and/or diarrhea, or very poor intake with dehydration associated with severe electrolyte disturbance, AKI etc resulting in admission and SARS-CoV-2 positive)

2.) **Possibly or likely COVID-19 associated admission:**
   a. Thromboembolic event (stroke, AMI, PE, DVT) during or shortly after SARS-CoV-2 infection: possible versus likely depends on presence or absence of predisposing

comorbidities (eg., history of stroke, AMI, PE, diabetes, smoking, BCP, etc) and member characteristics (e.g., age)

b. Cardiac dysrhythmia during or shortly after SARS-CoV-2 infection: probability depends on prior history of dysrhythmia, dysrhythmia-free interval prior to admission, etc.
c. Members infected with SARS-CoV-2 and another respiratory pathogen that may be associated with similar symptoms may be COVID-19 associated since it is not possible to determine the relative contribution of each pathogen to the symptoms.

**3.) Unlikely COVID-19 associated:**

a. incidentally SARS-CoV-2 positive admitted for an unrelated reason (e.g., L&D, elective surgery, baceterial infection such as abscess, cellulitis, etc) without evidence of COVID-19 associated symptoms that would have resulted in admission
b. admitted for a condition that may be COVID-19 associated (e.g., stroke, cardiac dysrhythmia) but has history of frequent strokes, underlying dysrhythmia, make it less likely that the admission is COVID-19 associated.

**Table S2. Risk of COVID-19 outcome among those who received a BNT162b2 XBB1.5-adapted vaccine by comparison and age group among adults ≥18 years of age**

| Comparison (reference) and age groups | Hospitalization | ED/UC encounter | Outpatient visit |
|---|---|---|---|
| | *Adjusted Odds ratio (95% CI)** | | |
| No XBB1.5-adapted vaccine | | | |
| ≥18 years | 0.37 (0.20–0.67) | 0.42 (0.34–0.53) | 0.42 (0.27–0.66) |
| 18-64 years | 0.32 (0.04–2.48) | 0.36 (0.24–0.54) | 0.68 (0.46–1.01) |
| ≥65 years | 0.37 (0.20–0.69) | 0.45 (0.34–0.59) | 0.32 (0.21–0.51) |
| BA.4/5-adapted bivalent vaccine but no XBB1.5-adapted vaccine | | | |
| ≥18 years | 0.40 (0.21–0.75) | 0.43 (0.34–0.55) | 0.49 (0.35–0.68) |
| 18-64 years | 0.35 (0.04–2.99) | 0.40 (0.26–0.62) | 0.78 (0.50–1.21) |
| ≥65 years | 0.39 (0.20–0.76) | 0.43 (0.31–0.58) | 0.29 (0.18–0.47) |
| ≥3 doses of wild-type vaccine but no variant-adapted vaccines of any kind (e.g., XBB1.5-adapted or BA.4/5-adapted or BA.1-adapted bivalent vaccines) | | | |
| ≥18 years | 0.36 (0.20–0.65) | 0.41 (0.33–0.51) | 0.44 (0.33–0.60) |
| 18-64 years | 0.27 (0.03–2.14) | 0.34 (0.23–0.51) | 0.60 (0.40–0.90) |
| ≥65 years | 0.36 (0.19–0.68) | 0.45 (0.34–0.60) | 0.35 (0.22–0.55) |
| ≥2 doses of wild-type vaccine but no variant-adapted vaccines of any kind (e.g., XBB1.5-adapted or BA.4/5-adapted or BA.1-adapted bivalent vaccines) | | | |
| ≥18 years | 0.37 (0.20–0.67) | 0.42 (0.33–0.52) | 0.46 (0.34–0.62) |
| 18-64 years | 0.30 (0.04–2.32) | 0.35 (0.23–0.52) | 0.65 (0.43–0.97) |
| ≥65 years | 0.37 (0.20–0.70) | 0.46 (0.35–0.61) | 0.33 (0.21–0.53) |
| Unvaccinated | | | |
| ≥18 years | 0.32 (0.16–0.64) | 0.40 (0.31–0.52) | 0.57 (0.39–0.84) |
| 18-64 years | 0.37 (0.04–3.22) | 0.37 (0.24–0.56) | 0.83 (0.52–1.33) |
| ≥65 years | 0.29 (0.14–0.61) | 0.33 (0.22–0.49) | 0.40 (0.18–0.87) |

*Models adjusted for week of encounter, age, sex, self-reported race/ethnicity, BMI, Charlson comorbidity index, prior SARS-CoV-2 infection, and utilization history (flu and pneumococcal vaccination, inpatient, ED, and outpatient encounters in prior year). BMI not included in hospitalization models for age <65 due to convergence issues.

**Table S3. Risk of COVID-19 outcome among those who received only prior (non-XBB1.5-adapted) vaccine doses compared to the unvaccinated among adults ≥18 years of age, by age group**

| **COVID-19 vaccination status (*vs* unvaccinated)** | **Hospitalization** | **ED/UC encounter** | **Outpatient visit** |
|---|---|---|---|
| | *Adjusted Odds ratio (95% CI)** | | |
| BA.4/5-adapted bivalent vaccine but no XBB1.5-adapted vaccine | | | |
| ≥18 years | 0.78 (0.50–1.20) | 0.92 (0.76–1.10) | 1.18 (0.88–1.56) |
| 18-64 years | 1.08 (0.41–2.90) | 0.90 (0.72–1.13) | 1.01 (0.73–1.41) |
| ≥65 years | 0.71 (0.43–1.17) | 0.77 (0.56–1.08) | 1.42 (0.71–2.83) |
| ≥3 doses of original wild-type vaccine but no variant-adapted vaccines of any kind (e.g., XBB1.5-adapted or BA.4/5-adapted or BA.1-adapted bivalent vaccines) | | | |
| ≥18 years | 0.89 (0.61–1.32) | 0.98 (0.84–1.14) | 1.36 (1.06–1.74) |
| 18-64 years | 1.34 (0.60–3.01) | 1.07 (0.89–1.27) | 1.38 (1.06–1.81) |
| ≥65 years | 0.79 (0.50–1.24) | 0.73 (0.54–0.99) | 1.15 (0.59–2.24) |
| ≥2 doses of wild-type vaccine but no variant-adapted vaccines of any kind (e.g., XBB1.5-adapted or BA.4/5-adapted or BA.1-adapted bivalent vaccines) | | | |
| ≥18 years | 0.84 (0.57–1.22) | 0.96 (0.83–1.11) | 1.24 (0.98–1.58) |
| 18-64 years | 1.21 (0.56–2.63) | 1.04 (0.88–1.22) | 1.22 (0.94–1.59) |
| ≥65 years | 0.74 (0.48–1.15) | 0.71 (0.53–0.96) | 1.23 (0.64–2.37) |

*Models adjusted for week of encounter, age, sex, self-reported race/ethnicity, BMI, Charlson comorbidity index, prior SARS-CoV-2 infection, and utilization history (flu and pneumococcal vaccination, inpatient, ED, and outpatient encounters in prior year). BMI not included in hospitalization models for age <65 due to convergence issues.

**Table S4. Sensitivity analyses of the risk of COVID-19 outcome among those who received a BNT162b2 XBB1.5-adapted vaccine by comparison group among adults ≥18 years of age, including those who received antiviral or monoclonal antibody treatment* in the 30 days prior to their COVID-19 encounter**

| Comparison (reference) group | Hospitalization | ED/UC encounter | Outpatient visit |
|---|---|---|---|
| | *Adjusted Odds ratio (95% CI)*** | | |
| No XBB1.5-adapted vaccine | 0.36 (0.20–0.66) | 0.44 (0.37–0.53) | 0.53 (0.41–0.68) |
| BA.4/5-adapted bivalent vaccine but no XBB1.5-adapted vaccine | 0.39 (0.21–0.72) | 0.44 (0.36–0.54) | 0.54 (0.41–0.72) |
| ≥3 doses of wild-type vaccine but no variant-adapted vaccines of any kind (e.g., XBB1.5-adapted or BA.4/5-adapted or BA.1-adapted bivalent vaccines) | 0.35 (0.19–0.63) | 0.43 (0.36–0.52) | 0.49 (0.38–0.64) |
| ≥2 doses of wild-type vaccine but no variant-adapted vaccines of any kind (e.g., XBB1.5-adapted or BA.4/5-adapted or BA.1-adapted bivalent vaccines) | 0.36 (0.20–0.65) | 0.44 (0.36–0.53) | 0.51 (0.39–0.67) |
| Unvaccinated | 0.32 (0.16–0.65) | 0.45 (0.36–0.57) | 0.73 (0.52–1.03) |

* Nirmatrelvir/ritonavir or any other COVID-19 outpatient antiviral or monoclonal antibody (i.e., molnupiravir, remdesivir, bebtelovimab, bamlanivimab, casirivimab, cilgavimab, sotrovimab, tixagevimab

**Models adjusted for week of encounter, age, sex, self-reported race/ethnicity, BMI, Charlson comorbidity index, prior SARS-CoV-2 infection, and utilization history (flu and pneumococcal vaccination, inpatient, ED, and outpatient encounters in prior year).
